# Molecular Feature-Based Classification of Retroperitoneal Liposarcoma: A Prospective Cohort Study

**DOI:** 10.1101/2024.08.09.24311657

**Authors:** Mengmeng Xiao, Xiangji Li, Fanqin Bu, Shixiang Ma, Xiaohan Yang, Jun Chen, Yu Zhao, Ferdinando Cananzi, Chenghua Luo, Li Min

## Abstract

**Background:** Retroperitoneal liposarcoma (RPLS) is a critical malignant disease with various clinical outcomes. However, the molecular heterogeneity of RPLS was poorly elucidated, and few biomarkers were proposed to monitor its progression.

**Methods:** RNA sequencing was performed on a training cohort of 88 RPLS patients to identify dysregulated genes and pathways using clusterprofiler. The GSVA algorithm was utilized to assess signaling pathways levels in each sample, and unsupervised clustering was employed to distinguish RPLS subtypes. Differentially expressed genes (DEGs) between RPLS subtypes were identified to construct a simplified dichotomous clustering via nonnegative matrix factorization. The feasibility of this classification was validated in a separate validation cohort (n=241) using immunohistochemistry (IHC) from the Retroperitoneal SArcoma Registry (RESAR). The study is registered with ClinicalTrials.gov under number NCT03838718.

**Results:** Cell cycle, DNA damage & repair, and Metabolism were identified as the most aberrant biological processes in RPLS, enabling the division of RPLS patients into two distinct subtypes with unique molecular signatures, tumor microenvironment, clinical features and outcomes (overall survival, OS and disease-free survival, DFS). A simplified RPLS classification based on representative biomarkers (LEP and PTTG1) demonstrated high accuracy (AUC>0.99), with patients classified as LEP+ and PTTG1- showing lower aggressive pathological composition ratio and fewer surgery times, along with better OS (HR=0.41, *P*<0.001) and DFS (HR=0.60, *P*=0.005).

**Conclusions:** Our study provided an ever-largest gene expression landscape of RPLS and established an IHC-based molecular classification that was clinically relevant and cost-effective for guiding treatment decisions.

## INTRODUCTION

Retroperitoneal liposarcoma (RPLS) is a soft tissue sarcoma (STS) originating in the retroperitoneum with an insidious onset. Traditional surgical resection has been regarded as a primary and curable treatment strategy of RPLS for the past fifty years (***Ecker et al., 2016***). However, the anatomical complexity and biological properties of sarcoma brought great difficulty to achieve microscopically margin-negative resection, leading to a high postoperative recurrence rate in RPLS patients. During the past decade, scientists tried to improve the postoperative survival of RPLS patients by personalized surgical resection and neoadjuvant/adjuvant therapies, but the effect was not satisfactory *(**Littau et al., 2020; Gronchi et al., 2015; Gronchi et al., 2009; Gronchi et al., 2013; Pisters et al., 2009***).

Recently, precision medicine greatly enriched the therapeutic approaches and reformed the clinical decision-making chain of tumor diagnosis and treatment, prolonging the median survival of main tumor types 2-10 times (***Kam et al., 2021; Zeng et al., 2022; Alifrangis et al., 2019; Frese et al., 2021***). Biomarker-based patient stratification and targeted therapy together make up the kernel of precision medicine, which is intrinsically based on the molecular profiling of cancers. However, our knowledge of the molecular features of RPLS is limited, and few clinically applicable molecular biomarkers and targeted drugs are available for RPLS treatment. Only sporadic molecules such as CDK4 (***Pilotti et al., 2000***), MDM2 (***Binh et al., 2005***), AURK4 (***Yen et al., 2019***) and CCNDBP1 (***Yang et al., 2021***) have been reported as prognostic and diagnostic biomarkers, but these biomarkers were poorly represented and verified. Therefore, it is crucial to reveal the molecular landscape of RPLS and explore a feasible classification for its diagnosis and treatment.

Here, we conducted a comprehensive inbestigation into the molecular characteristics of RPLS through the delineation of the largest gene expression landscape ever assembled for this rare disease entity. By identifying both RPLS-specific genes and prognostic biomarkers, we unveiled their intricate relationships with clinical parameters. Our findings revealed the existence of two distinct molecular subtypes within all RPLS patients, characterized by diverse pathological compositions, enriched signaling pathways, and varying clinical outcomes. This highlights the limitations of relying solely on traditional pathological classification for surgical decision-making in certain cases where patients exhibit favorable histological features but poor prognoses. Emphasizing the pivotal role of molecular subtyping in guiding individualized treatment strategies and enhancing patient management. To facilitate practical application in clinical settings, we developed a simplified RPLS classification system based on key biomarkers (LEP and PTTG1) representative of each subtype. Notably, this classification scheme was validated in a larger cohort of RPLS patients through immunohistochemistry assays (Figure 1), laying the groundwork for precise surgical interventions guided by molecular insights in the realm of RPLS treatment.

**Figure 1.**
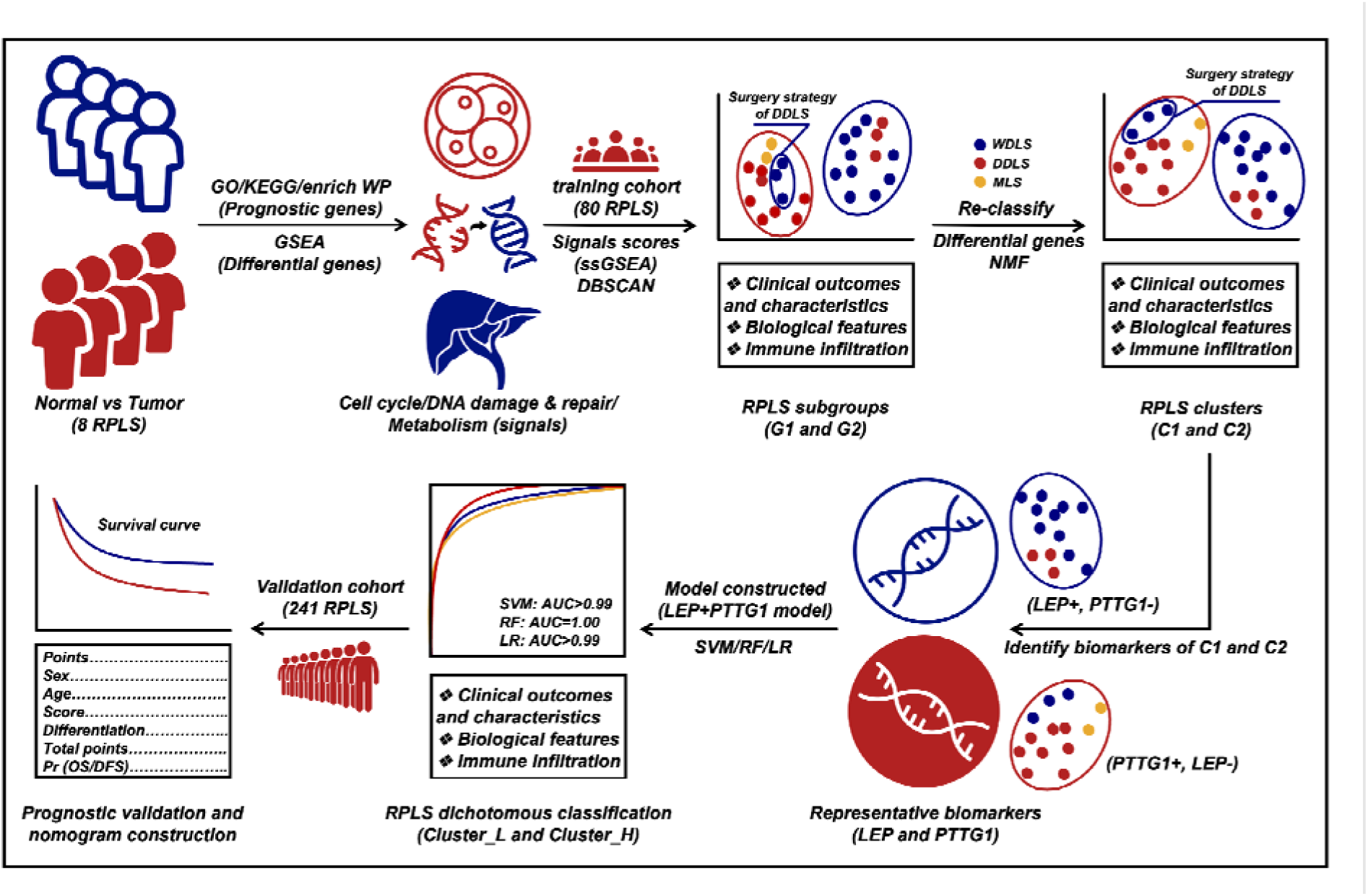
Flow diagram of exploring RPLS dichotomous classification

## MATERIALS AND METHODS

### Patients and tissue specimens

Patients who diagnosed with RPLS amenable to surgical resection were eligible for the study. The RPLS histology was confirmed according to the WHO criteria done on biopsy or surgical specimen by dedicated sarcoma pathologist. The exclusion criteria included the age<18 years; serious psychiatric disease that precludes informed consent or limits compliance; impossibility to ensure adequate follow-up. Tumor specimens from 88 RPLS patients (Training cohort 1, Table S1; Training cohort 2, Table S2) and another cohort of 241 RPLS patients (Validation cohort, Table S3) were obtained from our local Hospital. These cohorts are sourced from Retroperitoneal SArcoma Registry (RESAR, NCT03838718). All the patients underwent curative resection from January 2015 to May 2019. RPLS tissue specimens were snap-frozen in liquid nitrogen within 1 h and then stored in a -80 refrigerator before use. Clinical information was collected from the medical records, and no patient had undergone previous chemotherapy or radiotherapy. Overall survival (OS) was defined as the interval between the latest surgery and death from tumors or between the latest surgery and the last observation taken for surviving patients. Disease-free survival (DFS) was defined as the interval between the latest surgery and diagnosis of relapse or death. Informed consent for surgical procedures and specimen collection were obtained from each patient. This study has been reported in line with the REMARK criteria (***McShane et al., 2005***). The data of external validation cohort (GSE30929) was downloaded from GEO (https://www.ncbi.nlm.nih.gov/geo/).

### RNA sequencing, primary data processing, and analysis

Total RNA was extracted from Training cohort 1 (Table S4) and Training cohort 2 (Table S2) using TRIzol Reagent (Invitrogen). RNA degradation and contamination were monitored with 1% agarose gel. RNA purity was checked by the NanoPhotometer spectrophotometer (IMPLEN, Los Angeles, CA, USA). RNA concentration was measured using the Qubit RNA Assay Kit with the Qubit 2.0 Fluorometer (Life Technologies, CA, USA). RNA integrity was assessed using the RNA Nano 6000 Assay Kit of the Agilent Bioanalyzer 2100 System (Agilent Technologies, CA, USA).

A total amount of 3-5 ug RNA per sample was used as input material for the RNA library. Sequencing libraries were generated using NEBNext® Multiplex Small RNA Library Prep Set for Illumina® (NEB, USA) following the manufacturer’s recommendations and index codes were added to attribute sequences to each sample. The clustering of the index-coded samples was performed on a cBot Cluster Generation System using TruSeq SR Cluster Kit v3-cBot-HS (Illumia). After cluster generation, the strand-specific cDNA were sequenced on an Illumina NovaSeq 6000 platform, and single-end reads were generated (Novogene Bioinformatic Technology, Beijing, China).

FPKMs of mRNAs and non-coding RNAs in each sample were calculated by Cuffdiff (v2.1.1). FPKMs were calculated based on the length of the fragments and read counts mapped to this fragment. These sequencing data have been deposited at the Open Archive for Miscellaneous Data (OMIX) database of China National Center for Bioinformation (CNCB) under the accession number OMIX002786.

### Identification of differential genes

Gene difference analysis was performed to determine the differential genes (DEGs). An adjusted *FDR*<0.05 and |log2FC|>0.585 was considered significant. This process was conducted with the R package “limma”.

### Identification of prognostic genes

Cox univariate regression analysis was used to screen the prognostic genes of RPLS. Results of *P*<0.05 was considered significant. This process was conducted with the R package “survival”.

### Gene set enrichment analysis (GSEA) and immune infiltrate analysis

GSEA was performed in the tumor and normal groups to explore the biological signaling pathways. Pathway annotation files were downloaded from the msigdb (www.gsea-msigdb.org) platform. This process was conducted by the GSEA R package to elucidate the representative HALLMARK and REACTOME pathways enriched in RPLS patients. Immunocyte infiltration (immune score and stromal score) was measured by the Estimation of STromal and Immune cells in Malignant Tumor tissues using Expression data (ESTIMATE) algorithm. This process was completed via the “estimate” R package.

### Functional annotation

Functional enrichment analyses were performed to elucidate the possible biological processes and signaling pathways of the prognostic genes. Gene ontology (GO) and Kyoto Encyclopedia of Gene and Genomes (KEGG) analyses were conducted by R package “clusterprofiler”, and the false discovery rate<0.05 was considered significantly enriched.

### Consensus clustering with t-distributed stochastic neighbor embedding (t-SNE)

After evaluated the relative abundance level of related pathways, the Euclidean distance was calculated between any two samples and condensed into two-dimensional points using t-distributed stochastic neighbor embedding (t-SNE) (***Guo et al., 2019***) and subsequently visualized automatically with the density-based spatial clustering of applications with noise (DBSCAN) algorithm. This consensus clustering was conducted with the R packages “Rtsne” and “dbscan”.

### Consensus clustering with nonnegative matrix factorization (NMF)

Nonnegative matrix factorization (NMF) was used to perform RPLS subtyping. Specifically, NMF was applied to gene expression matrix *A* which contained gene sets of major signaling pathways and prognostic genes. Matrix *A* was factorized into 2 nonnegative matrices *W* and *H*. Repeated factorization of matrix *A* was performed and its outputs were aggregated to obtain consensus clustering of RPLS samples. The optimal number of subtypes was selected according to cophenetic, dispersion, and silhouette coefficients. This consensus clustering was conducted with the R package “NMF”.

### Construction of machine learning models

Machine learning models based on biomarkers were constructed by logistic regression (LR), support vector machine (SVM), and random forest (RF). These models were specifically tailored to analyze biomarker data in order to predict clinical outcomes in surgical patients.

LR is a statistical method that establishes a relationship between asset of independent variables and a binary outcome. It calculates the probability of an event occurring based on the input features derived from biomarkers relevant to surgical patient. SVM is a supervised learning algorithm that categorizes data points by identifying the optimal hyperplane that separates distinct classes within a high-dimensional space. This approach effectively maps biomarker data into a multidimensional space to facilitate accurate classification of patient outcomes. RF is an ensemble learning technique that generated multiple decision trees during training and aggregates the results to make predictions. By leveraging this method, we can enhance predictive accuracy by mitigating overfitting and increasing model robustness when analyzing biomarker-driven patient data.

The performance of these machine learning models was assessed using the area under the curve (AUC) metric. A higher AUC value indicates superior discriminatory power of model in distinguishing different clinical outcomes. An AUC value closer to 1.0 signifies strong predictive capability, while 0.5 indicates no discriminatory ability at all. By evaluating the AUC values generated by LR, SVM, and RF models, clinicians can identify which algorithm yields the most reliable predictions based on biomarker profiles for surgical patients.

### Immunohistochemistry

The protocol was performed as previously described (***Li et al., 2022***). In brief, the LEP and PTTG1 antibodies for immunohistochemistry were purchased from Proteintech (Cat No: bs- 0409R and bs-1881R). With deparaffinization for 15min × 3 in dimethylbenzene and routine hydration, the tissues were soaked in phosphate buffer saline (PBS) for 10min and then performed high-pressure antigen retrieval (Tris-EDTA, PH=9.0) for 2.5min. After being treated with a 3% endogenous catalase blocker (ZSBIO, PV-6000) for 10min, the tissues were incubated in goat serum (ZSBIO, ZLI-9022) for the blocking of nonspecific reaction and then incubated with primary antibody (LEP=1:300 and PTTG1=1:300) at 4 overnight. The next day, tissues were washed and incubated with goat anti-rabbit secondary antibody (ZSBIO, PV-9000) for 1h at room temperature, then washed and stained with DAB reagents (ZSBIO, ZLI-9018). Then hematoxylin staining, 1% hydrochloric acid alcohol differentiation, ammonia water anti-blue, and neutral gum sealing.

The IHC results were evaluated by pathologists, the staining extent was scored as 0-100%. The intensity score was defined as negative, low-expression, medium-expression, and high- expression, which were documented as 0, 1, 2, and 3 respectively. The final scores were calculated by the formula: *IHC score = Staining extent score* X *Staining intensity score*.

### Statistical Methods

R software (version 4.1.3) was used in this study. For quantitative variables, differences between the two groups and among multiple groups were analyzed by Wilcoxon’s test and One- way analysis of variance (ANOVA), respectively. For categoric variables, groups were compared by use of Chi-square test. Survival curves were determined by Log-rank test. The clinicopathological features and levels of immune infiltration were conducted by Wilcoxon’s test. A difference of *P*<0.05 indicated statistical significance unless specified otherwise.

## RESULTS

Baseline characteristics were shown in Table 1. Of 329 RPLS patients, 88 in training cohort and 241 in validation cohort. No statistically significant differences were found in the age, sex, pathology, surgery times, tumor size, and multilocation between the two cohorts (*P*>0.05). The IHC score of LEP and PTTG1 were 1.62 (0.820) and 0.830 (0.75) in validation cohort.

**Table 1.** Baseline characteristics of training cohort and validation cohort

|  | Training cohort (N=80)<sup>†</sup> | Validation cohort (N=241) | P value |
| --- | --- | --- | --- |
| Age (y) | 56.34 (11.14)<sup>‡</sup> | 55.11 (10.80)<sup>‡</sup> | 0.384 |
| Sex |  |  |  |
| Male | 37 (46.25) | 118 (48.96) | 0.674 |
| Female | 43 (53.75) | 123 (51.04) |  |
| Pathology |  |  |  |
| WDLs | 29 (36.25) | 75 (31.12) | 0.078 |
| DDLS | 48 (60.00) | 144 (59.75) |  |
| MLS and PLS | 3 (3.75) | 5 (2.07) |  |
| NR | 0 (0) | 17 (7.06) |  |
| Surgery times<sup>††</sup> |  |  |  |
| 0-1 | 50 (62.50) | 143 (59.34) | 0.834 |
| 2-3 | 21 (26.25) | 73 (30.29) |  |
| 4-7 | 9 (11.25) | 24 (9.96) |  |
| NR | 0 (0) | 1 (0.41) |  |
| Tumor size |  |  |  |
| All | 18.65 (8.70)<sup>‡</sup> | 16.90 (7.94)<sup>‡</sup> | 0.101 |
| <18 cm | 40 (50.00) | 135 (56.02) | 0.095 |
| >18 cm | 39 (48.75) | 92 (38.17) |  |
| NR | 1 (1.25) | 14 (5.81) |  |
| Multilocation |  |  |  |
| Yes | 52 (65.00) | 153 (63.49) | 0.081 |
| No | 28 (35.00) | 74 (30.71) |  |
| NR | 0 (0) | 14 (5.80) |  |
| MDM2 score |  |  |  |
| 0 | 11 (13.75) | NA | NA |
| 1 | 12 (15.00) | NA |  |
| 2 | 39 (48.75) | NA |  |
| 3 | 5 (6.25) | NA |  |
| 4 | 9 (11.25) | NA |  |
| NR | 4 (5.00) | NA |  |
| LEP score | NA | 1.62 (0.82)<sup>‡</sup> | NA |
| LEP strength |  |  |  |
| 0 | NA | 10 (4.15) | NA |
| 1 | NA | 39 (16.18) |  |
| 2 | NA | 74 (30.71) |  |
| 3 | NA | 115 (47.72) |  |
| NR | NA | 3 (1.24) |  |
| PTTG1 score | NA | 0.83 (0.75)<sup>‡</sup> | NA |
| PTTG1 strength |  |  |  |
| 0 | NA | 38 (15.77) | NA |
| 1 | NA | 100 (41.49) |  |
| 2 | NA | 61 (25.31) |  |
| 3 | NA | 39 (16.18) |  |
| NR | NA | 3 (1.25) |  |

<sup>†</sup>Clinical information missing in 8 RPLS patients (Training cohort 2)
<sup>‡</sup>The data is shown as Mean (SD); other data is shown as Number (%)
<sup>††</sup>The definition of surgical times is the sum of current admission surgery and previous surgical resection
DDLs, Dedifferentiated liposarcoma; LEP, Leptin; MDM2, Mouse double minute 2; MLS, Myxoid
liposarcoma; NA, Not applicable; NR, Not reported; PLS, Pleomorphic liposarcoma; PTTG1, Pituitary tumor
transforming gene 1; WDLs, Well-differentiated liposarcoma

### Cell cycle, DNA damage and repair, and Metabolism are dysregulated in RPLS

To reveal the general molecular features of RPLS compared to noncancerous adipose tissues, we first recruited 8 RPLS patients and collected paired tumor and normal tissues for differentially expressed gene (DEG) analysis. A total of 1354 DEGs, 554 upregulated and 800 downregulated, were identified (Figure 2A-B). To assess the underlying pathways of RPLS, GSEA analyses were performed for those DEGs. We found that proliferation-associated pathways, such as Mitotic spindle, E2F target, G2/M checkpoint and Separation of sister chromatids, were mainly enriched in tumors; while metabolism-related pathways, such as Bile acid metabolism, Heme and fatty acid metabolism and Integration of energy metabolism, were enriched in normal controls (Figure 2C).

**Figure 2.**
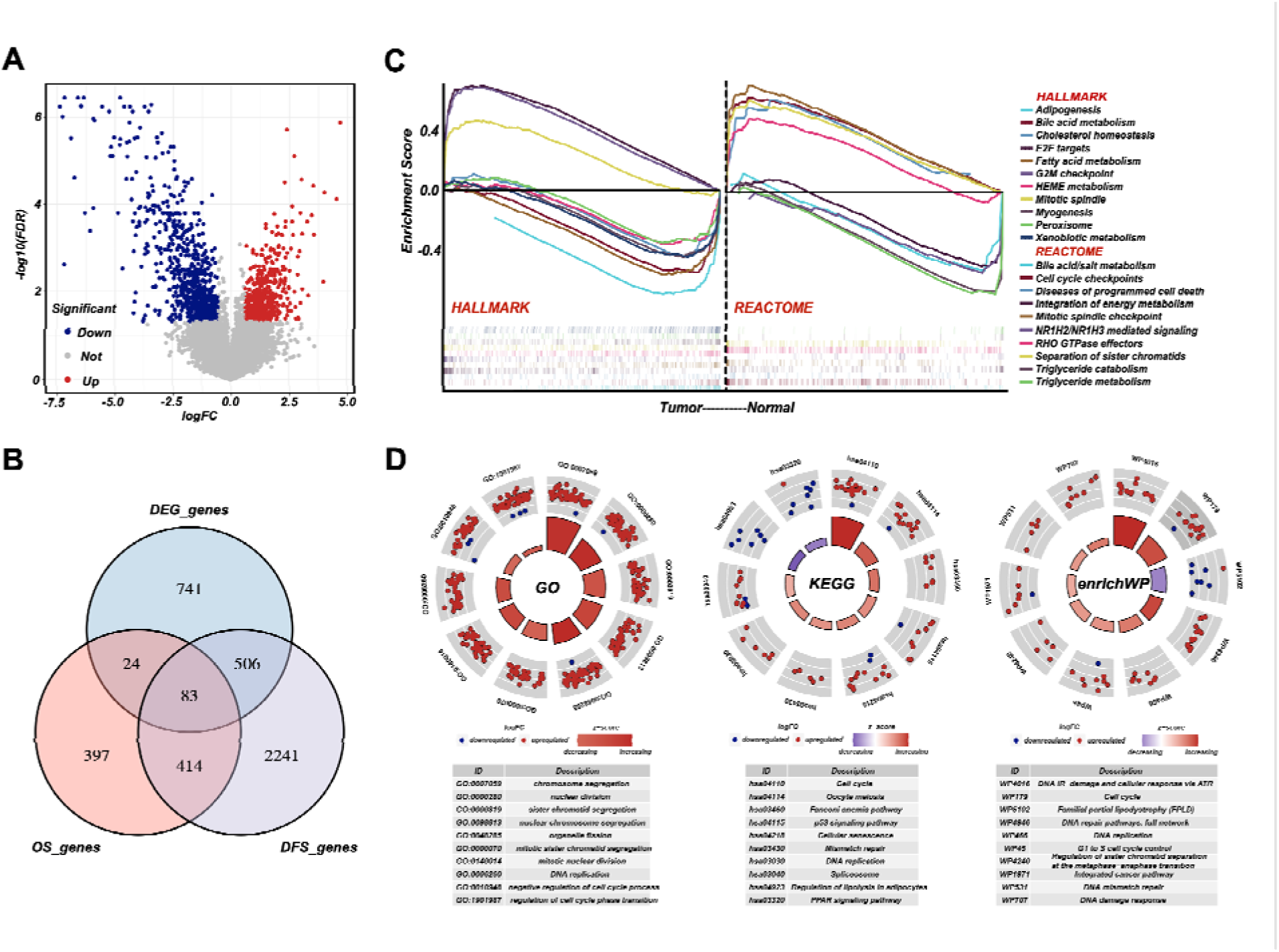
Cell cycle, DNA damage and repair, and metabolism are dysregulated in RPLS. Volcano plot of the DEGs in 8 normal vs 8 RPLS tissues **(A)**. Venn diagram showed shared genes between DEGs and prognostic genes **(B)**. GSEA analysis of RPLS tumors, including HALLMARK gene sets and REACTOME gene sets **(C)**. Circular plots of the prognostic genes in GO, KEGG, and enrichWP **(D)**.

Then, we collected another 80 samples to investigate the molecular heterogeneity of RPLS. Gene expression profiles showed 918 and 3244 genes associated with overall survival (OS) and disease-free survival (DFS), respectively. Among 497 candidate genes associated with both OS and DFS, 83 of them also overlapped with DEGs (Figure 2B). Functional annotation (GO, KEGG, and enrichWP) demonstrated that Cell cycle, DNA damage and repair, and Metabolism-related pathways were significantly enriched (Figure 2D), suggesting these signaling pathways were dysregulated in RPLS.

### RPLS subgroups based on molecular features show different clinical outcomes

To evaluate heterogeneous molecular clustering characteristics in RPLS, ssGSEA emerged as a widely adopted method for computing the enrichment level of specific biological signaling pathways for each sample based on gene expression data. This aids in gaining insights into the overall activity level of signaling pathways. Here, we scored each sample on the dysregulated pathways by ssGSEA and divided RPLS patients into two subgroups (Figure 3A). Subgroup 1 (G1) showed better OS and DFS compared to subgroup 2 (G2) (Figure 3B-C), G1 displayed elevated ssGSEA scores associated with Metabolism, whereas G2 exhibited heightened ssGSEA scores linked on Cell cycle and DNA damage and repair (Figure 3D). These features suggested that effective monitoring of the prognosis of RPLS patients can be achieved based on the activation status of specific pathways. We also evaluated the clinical features and immune infiltration levels of those samples. The results showed that G1 had lower aggressive pathological composition ratio (Figure 3E), MDM2 (Figure 3G) and Ki67 (Figure S1A) expression, larger tumor size (Figure S1B), and higher tumor microenvironment (TME) level compared to G2 (Figure 3H).

**Figure 3.**
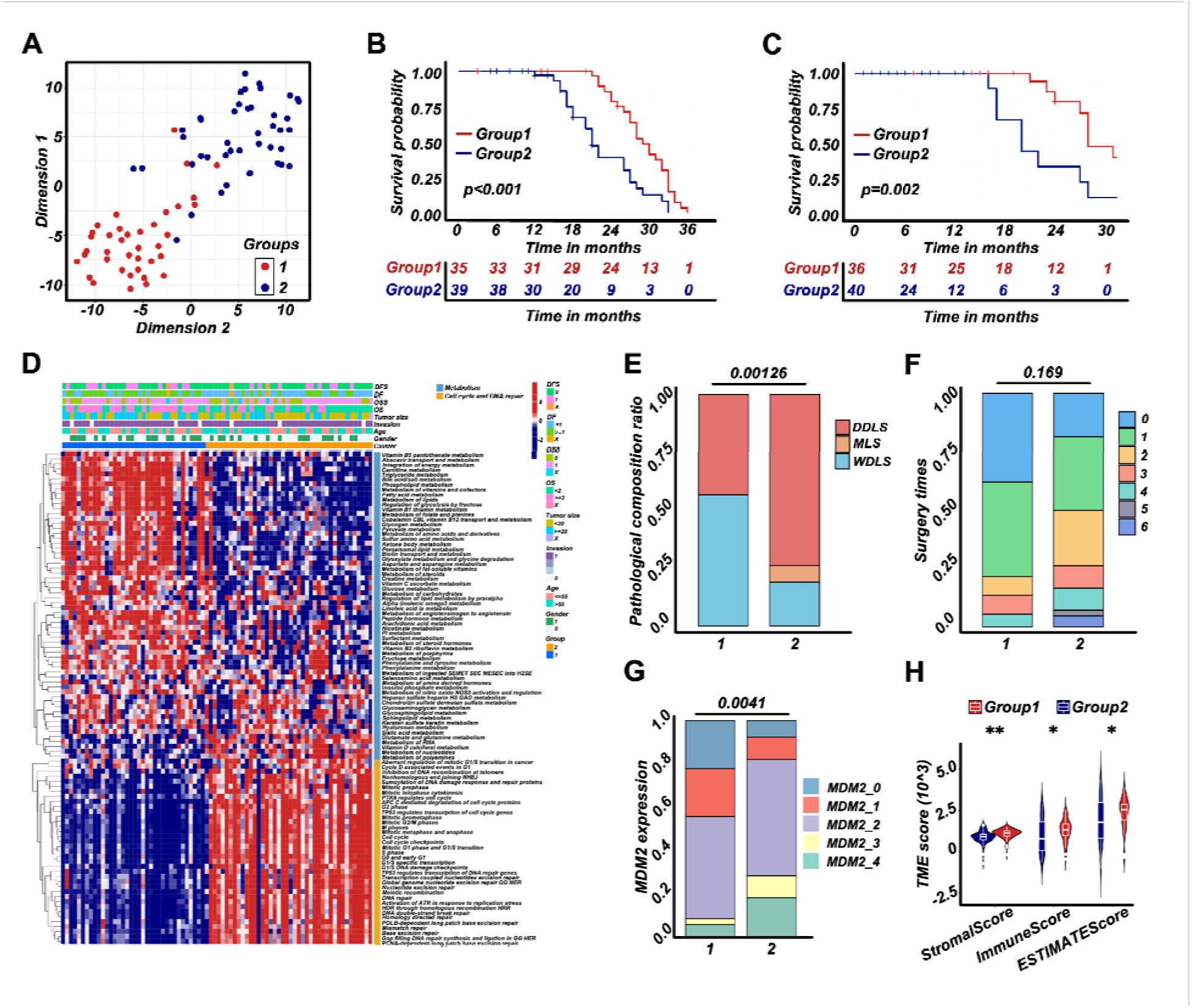
RPLS subgroups (G1 and G2) based on cell cycle, DNA damage and repair, and metabolism tSNE exhibited the subgroups (G1 and G2) of RPLS **(A)**. Survival cures of OS **(B)** and DFS **(C)** in G1 and G2. The hierarchical clustering heatmap of dysregulated pathways in G1 and G2 **(D)**. Histograms revealed the difference of pathological composition ratio **(E)**, surgery times **(F)**, and MDM2 **(G)** in G1 and G2. Violin plot of the microenvironmental scores in G1 and G2 **(H)**.

Surgery times for G1 were also tended to decrease (Figure 3F). Taken together, the above restuls indicated that RPLS subgroups based on molecular features showed distinct clinical features and clinical outcomes.

### A simplified RPLS classification strategy derived from RPLS G1/G2 subgroups

To explore representative biomarkers for different RPLS subgroups, we performed a DEG analysis between G1 and G2. There were 1258 genes downregulated, among which 112 of them indicated good prognosis (protective genes). Correspondingly, 754 genes were upregulated, and 28 of them indicated poor prognosis (aggressive genes) (Figure S1C-D). Enrichment analysis suggested that those DEGs were also associated with cell cycle regulation and metabolism (Figure S1E), which was consistent with previous results (Figure 2C).

To develop a simplified RPLS clustering based on DEGs, we adopted NMF and tSNE for a re-classification of those patients. The results showed RPLS patients were also divided into two clusters (Figure 4A and Figure S1F-G). We then annotated the samples of two clusters by ssGSEA and found Cluster1 (C1) was related to metabolic processes, and Cluster2 (C2) was mainly related to the processes of Cell cycle and DNA damage and repair (Figure S2A). Also, C1 showed better OS and DFS, lower pathological composition ratio and MDM2 expression, and fewer surgery times (Figure 4B-F). Lower Ki67 expression and larger tumor size were observed in C2 (Figure S2B-C). Interestingly, the biological annotations of the C1/C2 classification were greatly consistent with G1/G2. Therefore, a simplified RPLS classification strategy derived from RPLS subgroups was provisionally established.

**Figure 4.**
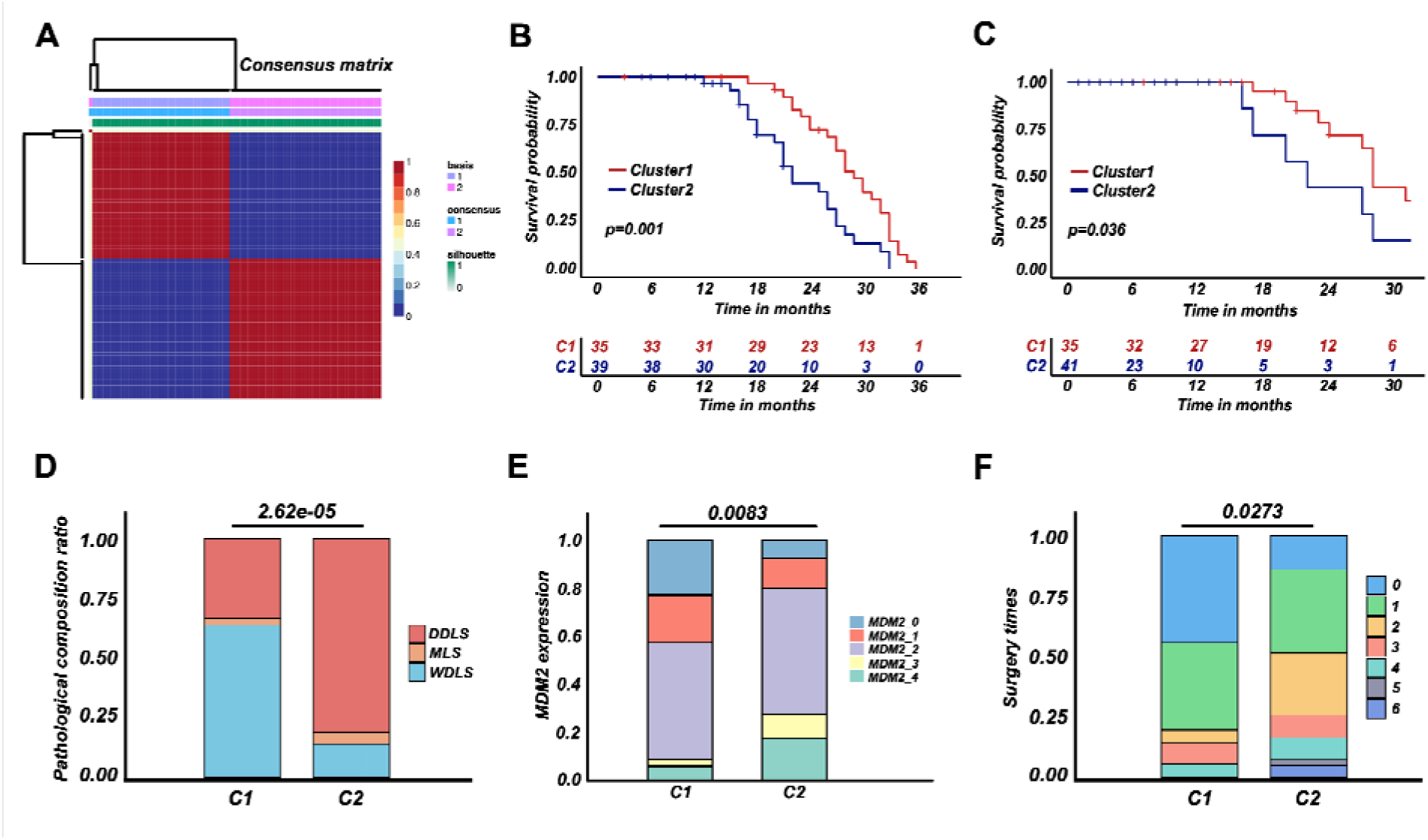
RPLS classification strategy (C1 and C2) derived from RPLS subgroups NMF for a re-classification of training cohort 1 (C1 and C2) **(A)**. Survival cures of OS **(B)** and DFS **(C)** in C1 and C2. Histograms revealed the difference in pathological composition ratio **(D)**, MDM2 **(E)**, and surgery times **(F)** in C1 and C2.

### Development of a dichotomous RPLS classification model

For NMF classification of RPLS patients, LEP and PTTG1 were identified as representative biomarkers of C1 and C2, respectively (Figure 5A). The selection criteria for identifying LEP and PTTG1 as biomarkers involved selecting prognostic genes that were highly expressed in C1 and C2, respectively, and achieved the highest AUC value in distinguishing the two RPLS groups. We aimed to replicate the RPLS classification of C1 and C2 by integrating these two biomarkers with the assistance of machine learning algorithms, and this two-gene panel achieved promising results (Logistic, AUC=0.995; SVM, AUC=0.997; RF, AUC=1.000; Figure 5B). Also, a linear negative correlation between LEP and PTTG1 expression was detected (Figure 5C).

**Figure 5.**
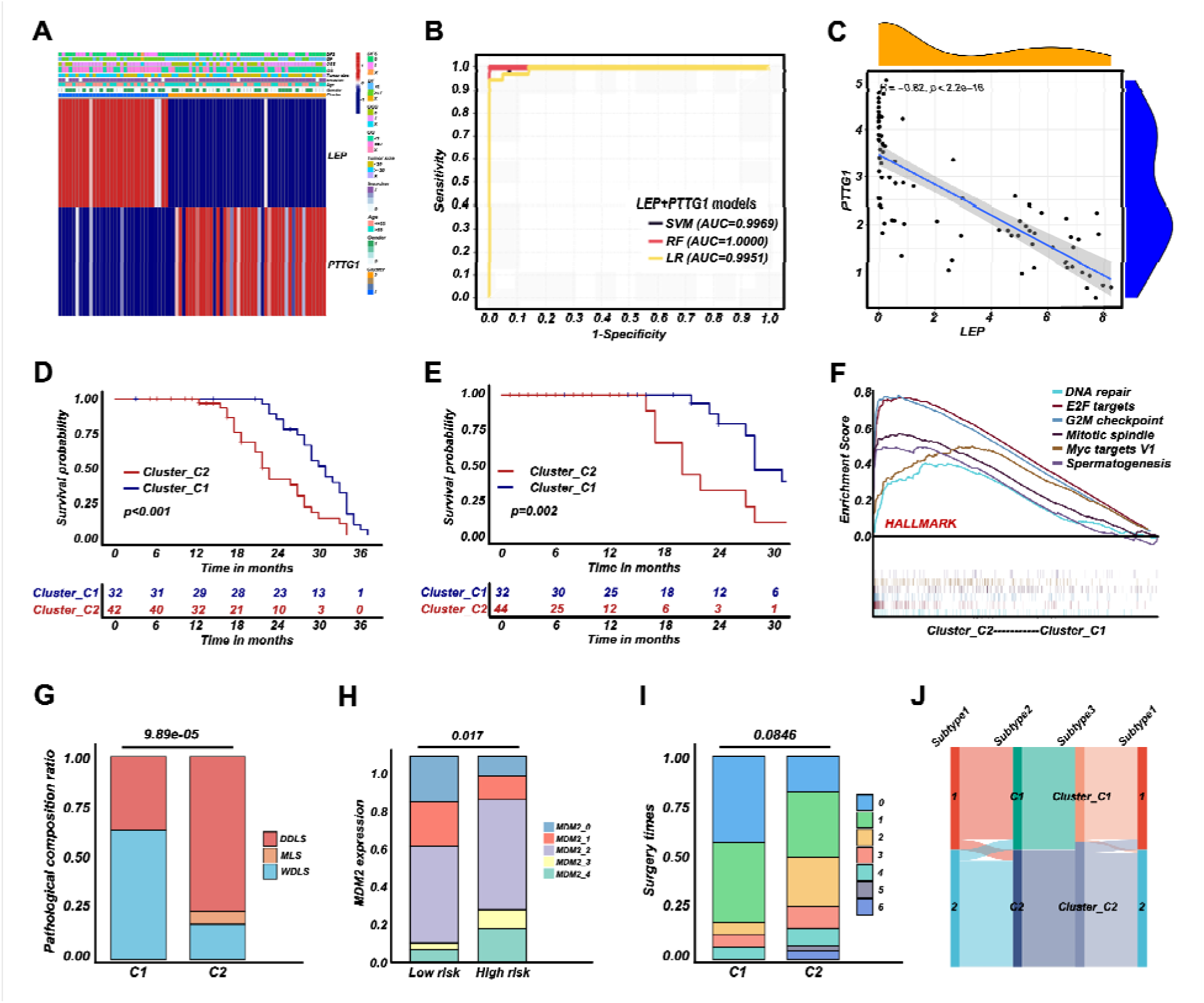
RPLS dichotomous classification (Cluster_C1 and Cluster_C2) derived from RPLS clusters Heatmap of biomarkers identified (LEP and PTTG1) in C1 and C2 **(A)**. ROC curves of the machine learning models to identify C1 and C2 **(B)**. Correlation between LEP and PTTG1 expression **(C)**. Survival curves of OS **(D)** and DFS **(E)** in Cluster_C1 (low risk) and Cluster_C2 (high risk) groups. GSEA of HALLMARK gene sets in Cluster_C1 and Cluster_C2 **(F)**. Histograms revealed the difference of pathological composition ratio **(G)**, MDM2 level **(H)**, and surgery times **(I)** in Cluster_C1 and Cluster_C2. Sankey diagram indicated the correlation among G1/G2, C1/C2, and Cluster_C1/C2 **(J)**.

Considering the enhanced interpretability and generalization of linear models, we adopted the results of Logistic regression for subsequent analysis (Risk values=2.182×PTTG1-2.204×LEP). The patients marked as high-risk (Cluster_H) exhibited worse OS and DFS than those marked as low-risk (Cluster_L) (Figure 5D-E). Dysregulated pathways, such as DNA repair and Cell cycle regulation, were enriched in Cluster_H (Figure 5F and Figure S2D), and Cluster_H presented more aggressive pathological composition ratio, higher MDM2 levels and marginally increased in surgery times than Cluster_L (Figure 5G-I). Similarly, the Cluster_H showed higher Ki67 level and smaller tumor sizes (Figure S2E-F). Moreover, a Sankey diagram was drawn to show the correlation among G1/G2, C1/C2, and Cluster_L/H. Cluster_L/H were well-matched to C1/C2 and G1/G2, suggesting LEP and PTTG1 were promising biomarkers for a dichotomous RPLS classification (Figure 5J). To ensure the broader applicability of LEP and PTTG1 as classification biomarkers, we performed an independent validation using an external liposarcoma cohort (GSE30929). This dataset was selected due to its relevance to RPLS (N=63, 45%) and the availability of distant recurrence-free survival (DRFS) outcomes, aligning with the clinical focus of our study. Applying our established logistic regression, we found that the high-risk (V) group exhibited significantly worse DRFS compared to the low-risk (V) group (Figure S3A-B), and the high-risk group (V) demonstrated a higher proportion of high-grade histology (Figure S3C-D), consistent with our previously findings. These results validate the robustness and generalizability of our risk stratification model across distinct liposarcoma cohorts. The external dataset’s alignment with our findings underscores the potential of LEP and PTTG1 as reproducible biomarkers for prognosis and therapeutic stratification in liposarcoma.

### Validation of the dichotomous RPLS classification in another 241 RPLS patients

To validate LEP and PTTG1 as biomarkers for a dichotomous RPLS classification, we performed IHC staining of two biomarkers in validation cohort. The representative images of LEP and PTTG1 with different expression levels were shown in Figure 6A-B. The IHC scores were integrated with the previously fitted coefficients to evaluate the prognosis of RPLS patients (Risk values=2.182×PTTG1_IHC_-2.204×LEP_IHC_). The cutoff value of validation cohort is the median of risk value. The high-risk group had worse OS and DFS (Figure 6C-D), along with more surgery times and more aggressive pathological composition ratio (Figure 7A-B), but the difference of tumor size was not observed between the two groups (Figure 7C). Then we constructed visual nomograms for a precise survival prediction of RPLS patients by combining the risk score with clinical features. The predictive abilities of the 1-, 2-, and 3-year OS (Figure 7D-F) and DFS (Figure S4A-C) were 0.743-0.788. Together, we proposed a simple and clinically applicable molecular classification strategy for RPLS patients.

**Figure 6.**
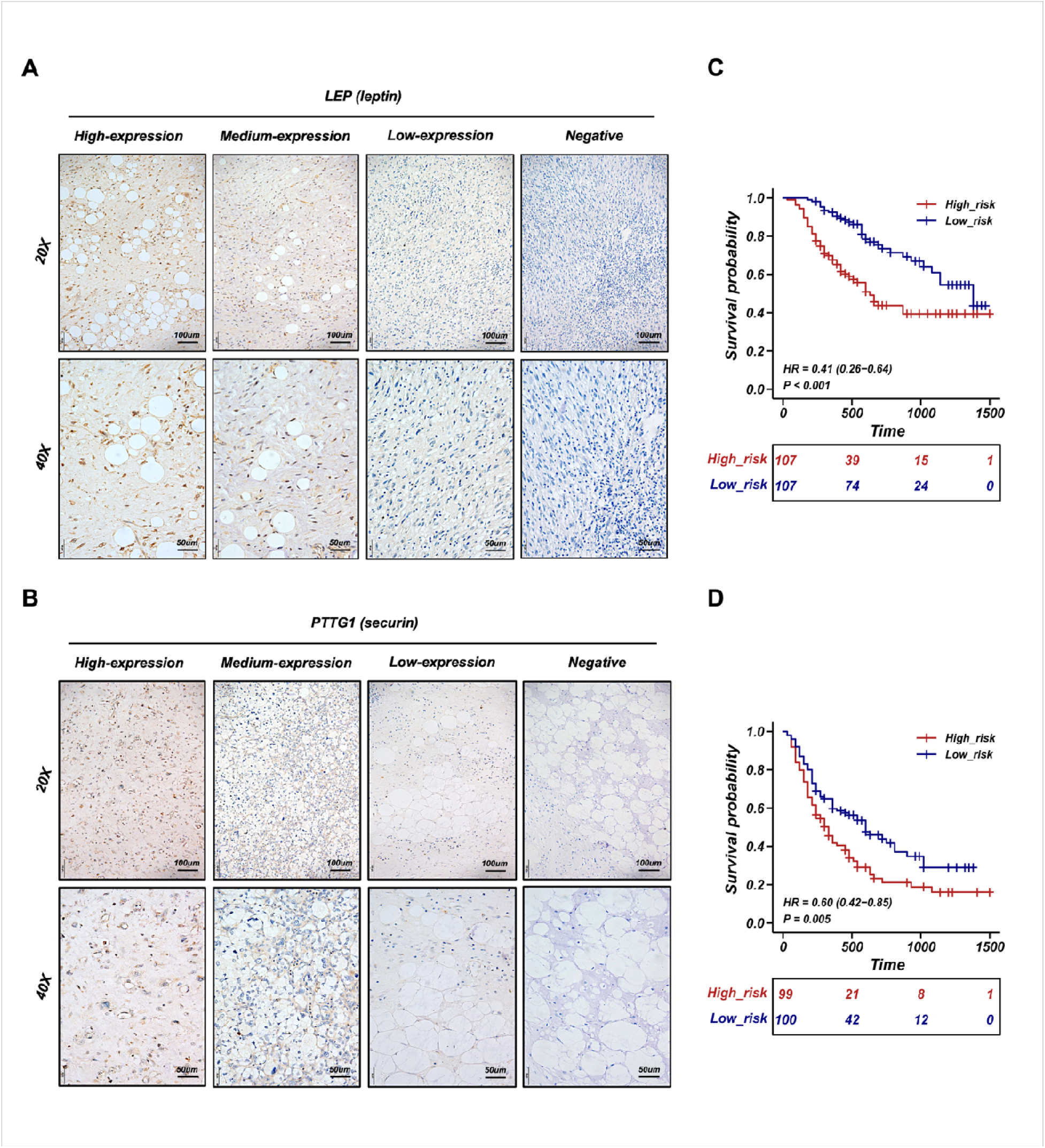
Validation of the RPLS dichotomous classification in another 241 RPLS cohort. Representative IHC staining images of LEP **(A)** and PTTG1 **(B)**. Survival curves of OS **(C)** and DFS **(D)** in high-risk and low-risk groups.

**Figure 7.**
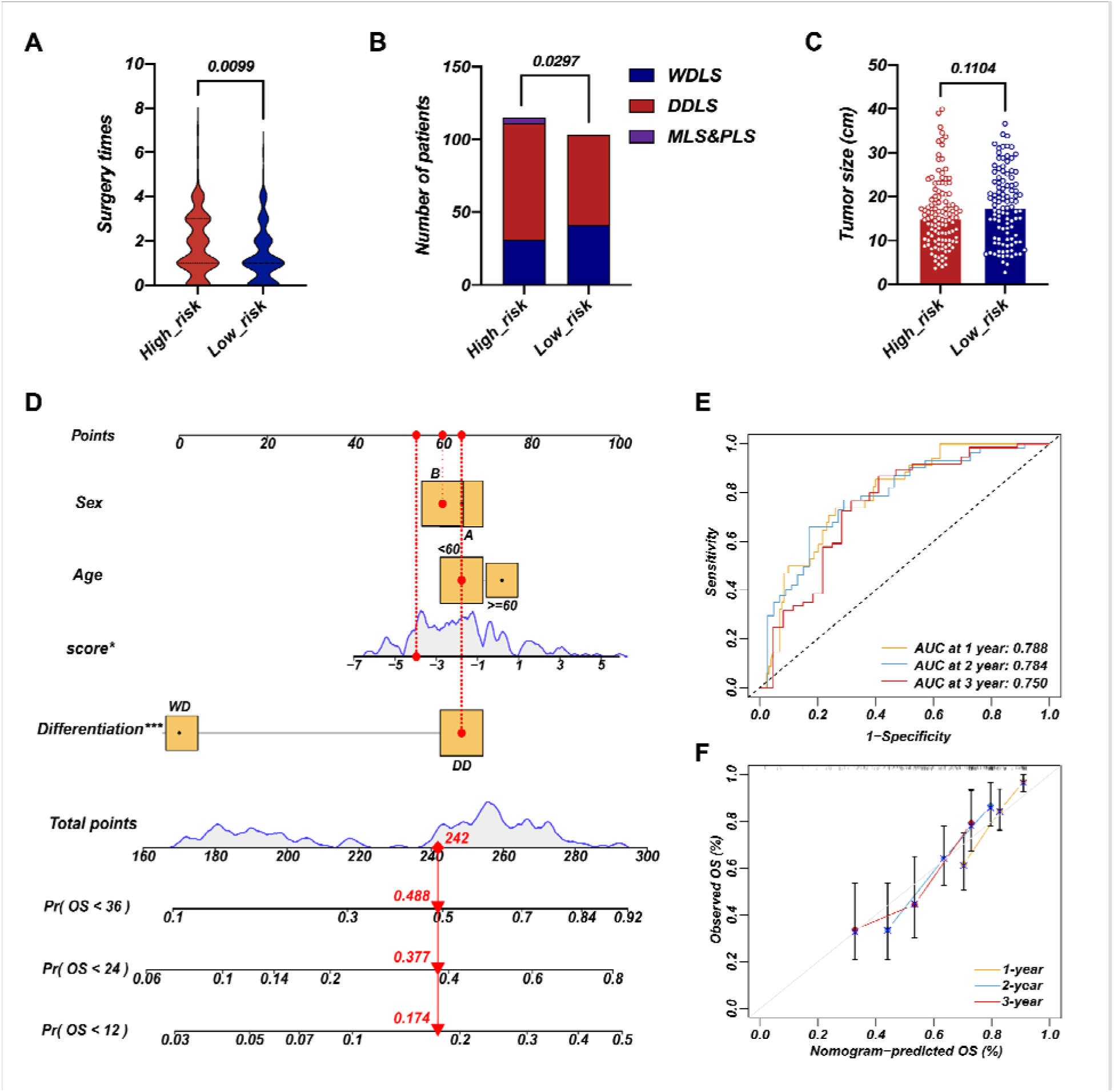
Survival nomogram of LEP+PTTG1 model in validation cohort The difference of surgery times **(A)**, pathological composition **(B)**, and surgery times **(C)** in high-risk and low-risk groups. Nomograms for OS was developed in REASR cohort with four factors: sex, age, risk score, and differentiation **(D)**. ROC curves of 1-, 2-, and 3-year OS in validation cohort **(E)**. Calibration curves of predicting 1-, 2-, and 3-year OS in validation cohort **(F)**.

## DISCUSSION

Here we divided RPLS patients into two subgroups based on Cell cycle, DNA damage & repair, and Metabolism-related pathways. G1 was annotated as Metabolism-active, which exhibited high ssGSEA scores on Metabolism-associated pathways, while G2 showed high ssGSEA scores on Cell cycle and DNA damage and repair with a high Ki67 and MDM2 level. had more aggressive molecular features and worse clinical outcomes compared to G1, in accordance with previously reported tumor classification (***Lindskrog et al., 2021; Yu et al., 2021; Zhang et al., 2022***). In fact, ***Demicco et al., 2017*** has integrated SCNA and DNA methylation divide dedifferentiated liposarcoma into two subtypes (S1 and S2), the unfavorable cluster was characterized as JUN amplified (An oncogene that promotes proliferation and metastasis) and lower inferred fraction of immature dendritic cells. However, the patients in ***Demicco et al., 2017*** were of complex origin (mixed limbs, trunk and retroperitoneum), providing limited guidance for RPLS molecular classification. Here we reported the first clinically applicable RPLS molecular classification based on RNA sequencing and IHC validation cohorts.

To facilitate the clinical application, we constructed a simplified RPLS molecular classification derived from the original Cell cycle/Metabolism subgroups. By NMF algorithm, we identified LEP and PTTG1 as representative biomarkers for each subtype. A model based on IHC staining of LEP and PTTG1 successfully approximated the original dichotomous RPLS classification in biological features and survival outcomes. LEP is an important regulator of basal metabolism and food intake, which is considered a linkage between metabolism and the immune system (***Jiménez-Cortegana et al., 2021***). Although LEP-based targeting therapies have not yet been fully applied, LEP has already been identified as a potent metabolic reprogramming agent to support antitumor responses in aggressive melanomas (***Waldman et al., 2020; De la et al., 2019; Rivadeneira et al., 2019***). In addition, LEP improves the immunotherapeutic effects by regulating innate and adaptive immune responses via increasing the cytotoxicity of NK cells (***Vera et al., 2018***), stimulating the proliferation of T/B cells (***Vera et al., 2018; Bernotiene et al., 2006***), and activating DC cells (***Hu et al., 2019***). Those reported roles of LEP provided a good mechanism explanation on the features of metabolism pathway-enriched, better prognosis, higher TME level of Metabolism subgroup (LEP+). In contrast, PTTG1 acts as a regulator of sister chromatid separation during cell division under physiological conditions (Zou et al., 1999), which is closely linked to genetic instability, aneuploidy, tumor progression, invasion, and metastasis (***Heaney et al., 2000; Ramaswamy et al., 2003; Kim et al., 2005; Yu et al., 2003; Teveroni et al., 2021; Romero et al., 2001***). PTTG1 also regulates the cell cycle and the transactivation of growth factors as an initiator and promoter of tumorigenesis (***Zou et al., 1999; Mora-Santos et al., 2013; McCabe et al., 2002; Ishikawa et al 2001; Hamid et al., 2005***). Overexpressing PTTG1 was correlated with worse prognosis in tumors, such as ovarian cancer (***Parte et al., 2019***), cervical cancer (***Guo et al., 2019***), renal cell carcinoma (***Tian et al., 2022***), and colorectal cancer (***Heaney et al., 2000***). Therefore, the biological functions of PTTG1 provided a good mechanism explanation of the pathway-enriched of cell cycle/DNA damage & repair-associated, worse prognosis, and more aggressive pathological composition ratio the Cell cycle subgroup (PTTG1+).

## CONCLUSION

Our study presented a comprehensive gene expression landscape of RPLS, revealing distinct molecular features. Through categorizing RPLS into Metabolism and Cell Cycle subtypes and identifying key biomarkers LEP and PTTG1, we established a dichotomous classification system verified by IHC assays. This innovative approach enables precise guidance for surgeons in adjusting treatment strategies for patients with histologically favorable but prognostically challenging RPLS cases, thereby advancing the implementation of precision medicine in guiding surgical interventions for RPLS.

## Data Availability

All data produced in the present study are available upon reasonable request to the authors

## Abbreviations

RPLS: Retroperitoneal liposarcoma
DEGs: Differentially expressed genes
IHC: Immunohistochemistry
STS: Soft tissue sarcoma
OS: Overall survival
DFS: Disease-free survival
OMIX: Open archive for miscellaneous data
CNCB: China national center for bioinformation
GSEA: Gene set enrichment analysis
GO: Gene ontology
KEGG: Kyoto encyclopedia of gene and genomes
t-SNE: t-distributed stochastic neighbor embedding
DBSCAN: Density-based spatial clustering of applications with noise
NMF: Nonnegative matrix factorization
LR: Logistic regression
SVM: Support vector machine
RF: Random forest
AUC: Area under curve

## Authors’ contributors

Mengmeng Xiao: Methodology, Formal analysis, Data curation, Writing-original draft, Writing-review & editing. Xiang Ji Li: Methodology, Formal analysis, Investigation, Data curation, Writing-original draft. Fanqin Bu: Conceptualization, Methodology, Formal analysis, Investigation, Data curation, Writing-review & editing. Shixiang Ma: Methodology, Formal analysis. Xiaohan Yang: Methodology, Investigation. Jun Chen: Formal analysis. Yu Zhao: Investigation. Ferdinando Cananzi: Investigation. Chenghua Luo: Conceptualization, Methodology, Writing-review & editing, Supervision. Li Min: Conceptualization, Resources, Writing-review & editing, Supervision.

## Ethics approval and consent to participate

Specimens of RPLS were obtained from Peking University International Hospital. The study protocol was approved by the Ethics Committee of Peking University International Hospital, Peking University Health Science Center (WA2020RW29) and conducted in accordance with Helsinki Declaration. All patients signed the informed consent.

## Finding information

This work was supported by grants from the Beijing Municipal Science and Technology Project (Z191100006619081), National Natural Science Foundation of China (82073390), and Young Elite Scientists Sponsorship Program (2023QNRC001). The study sponsors had no role in the design and preparation of this manuscript.

## Research registration unique identifying number (UIN)

1. Name of the registry: ClinicalTrials.gov
2. Unique identifying number or registration ID: NCT03838718
3. Hyperlink to your specific registration: https://clinicaltrials.gov

## Consent for publication

All authors have read and approved the manuscript and agree with submission to ***eLife***.

## Data availability

Data supporting the conclusions of this article are presented within the article and its supplementary files.

## Data anonymization

All participant identifiers were replace with unique IDs during data collection and analysis.

## Competing interests

The authors declare that they have no competing interests.

## Supplementary Information

Table S1. The detailed clinicopathological characteristics of the training cohort 1

Table S2. The RNA-seq data of the training cohort 2

Table S3. The detailed clinicopathological characteristics of the validation cohort

Table S4. The RNA-seq data of the training cohort 1

Figure S1. Clinical features and re-classification of RPLS subgroups (G1 and G2) The difference of Ki67 **(A)** and tumor size **(B)** in G1 and G2. Volcano plot of the DEGs (G1 vs G2) **(C)**. Venn diagram showed shared genes between DEGs and prognostic genes **(D)**. Bubble plot of the DEGs enrichment **(E)**. NMF for a re-classification of training cohort 1 **(F)**. tSNE exhibits the RPLS clusters (C1 and C2) **(G)**.

Figure S2. Dysregulated pathways and clinical features of RPLS clusters and high-/low-risk groups The hierarchical clustering heatmap of dysregulated pathways in C1 and C2 **(A)**. The difference of Ki67 **(B)** and tumor size **(C)** in C1 and C2. The hierarchical clustering heatmap of dysregulated pathways in high- and low-risk groups **(D)**. The difference of tumor size **(E)** and Ki67 **(F)** in high- and low-risk groups.

Figure S3. Validation of LEP+PTTG1 model in an external liposarcoma cohort Survival curve of DRFS in low-risk and high-risk groups **(A)**. ROC curves of 1-, 3-, and 5-year DRFS in validation cohort **(B).** Histograms revealed the difference of pathological composition ratio in low-risk and high-risk groups **(C)**. Pathological type details of LPS patietns **(D)**.

Figure S4. Survival nomogram of LEP+PTTG1 model in validation cohort Nomograms for DFS was developed in REASR cohort with four factors: sex, age, risk score, and differentiation **(A)**. ROC curves of 1-, 2-, and 3-year DFS in validation cohort **(B)**. Calibration curves of predicting 1-, 2-, and 3-year DFS **(C)**.

## Notes

### Competing Interest Statement

The authors have declared no competing interest.

### Summary of Updates

MATERIALS AND METHODS (Patients and tissue specimens) updated. Section on RESULTS (Development of dichotomous RPLS classification model) updated to clarify the selection criteria for identifying LEP and PTTG1. Section on RESULTS (Development of dichotomous RPLS classification model) updated to validate the broader applicability of LEP and PTTG1 as classification biomarkers. Supplemental files updated (added Figure S3). Figure legends of Figure S3 and Figure S4 updated.

