## Supplementary figures and images for "Molecular Feature-Based Classification of Retroperitoneal Liposarcoma: A Prospective Cohort Study"

### Figure S1.tiff

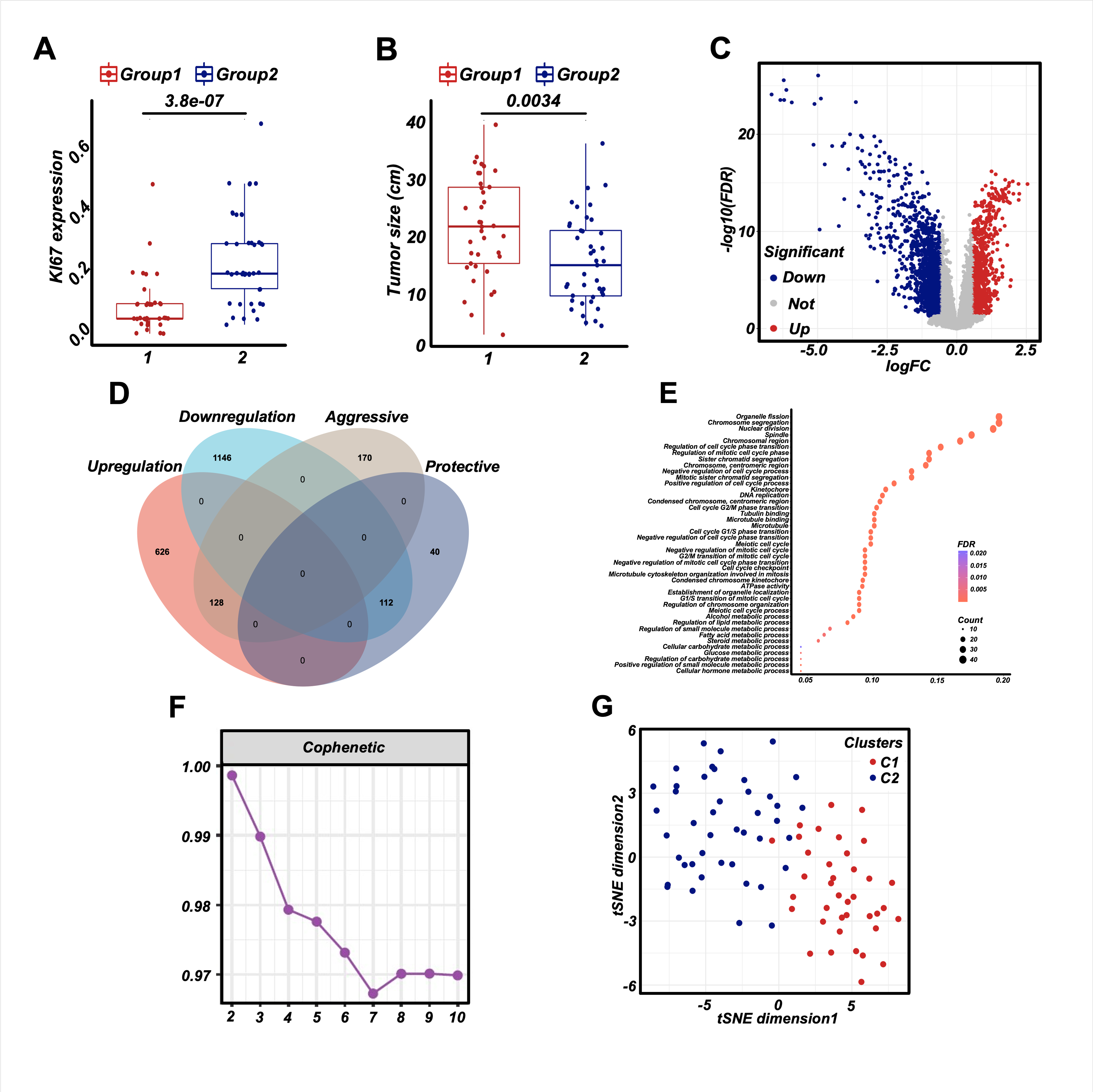

### Figure S2.tiff

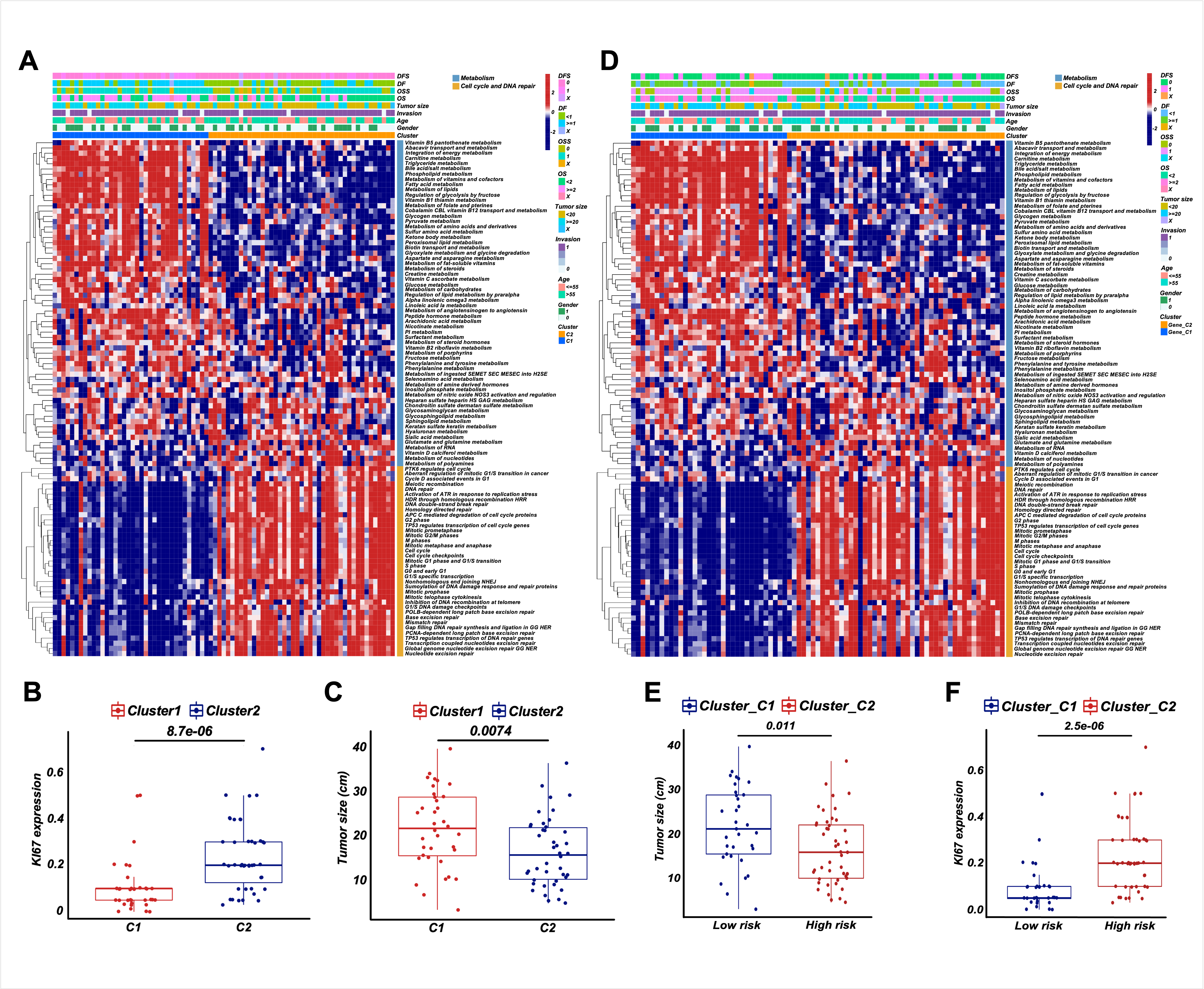

### Figure S3.tiff

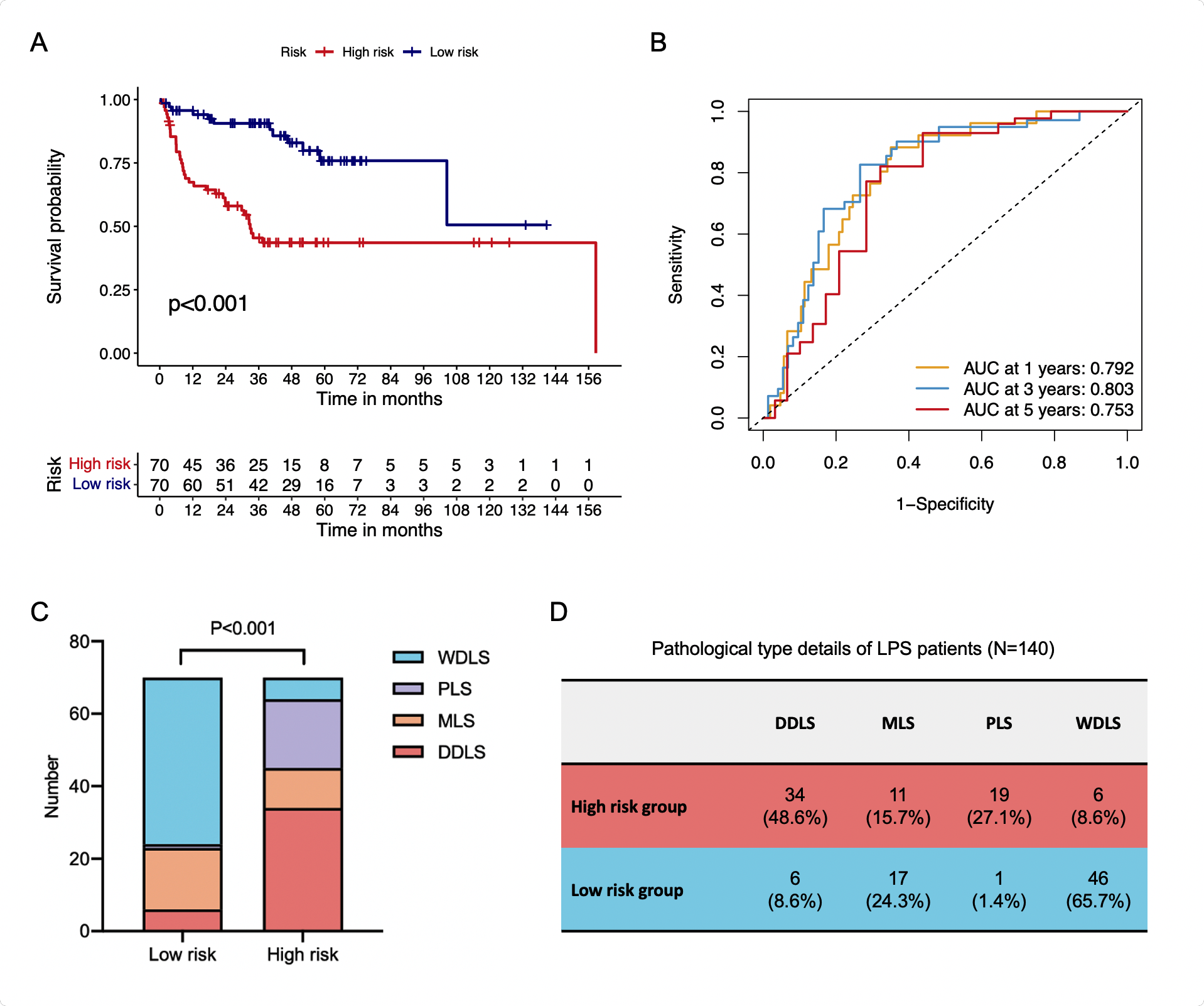

### Figure S4.tiff

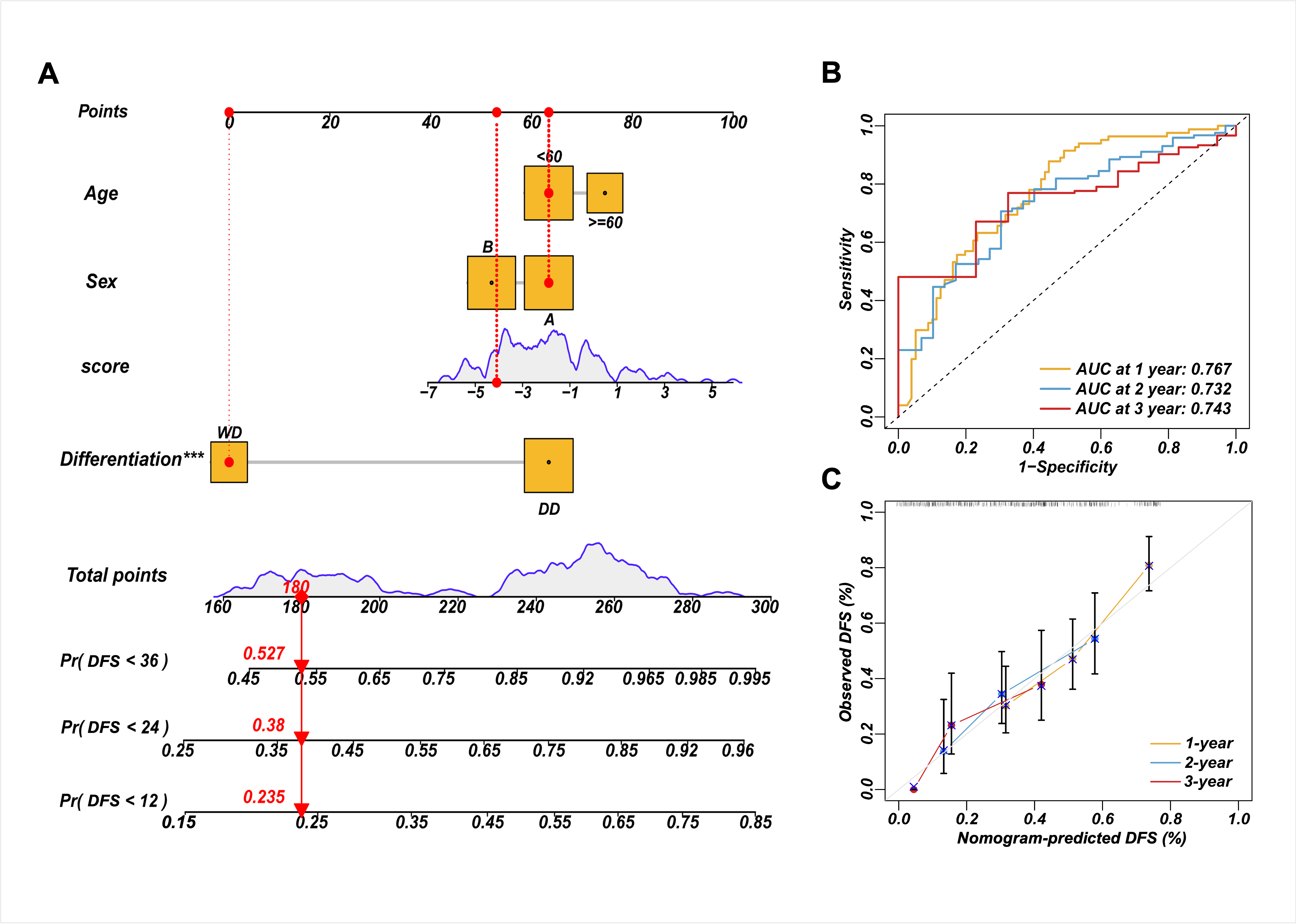
